# Effect of neonatal BCG vaccination on the evolution of Hashimoto’s Thyroiditis

**DOI:** 10.1101/2024.02.28.24303481

**Authors:** Kanhaiya Agrawal, Preetam Basak, Darshan Badal, Pinaki Dutta, Rama Walia, Naresh Sachdeva

**Author notes:** Corresponding Author: Prof. Naresh Sachdeva, Department of Endocrinology, Post Graduate Institute of Medical Education and Research (PGIMER), Chandigarh, INDIA, E mail. **Authors’ Contributions: KA** collected and processed all blood samples, performed all the experiments, managed the patients, collected and interpreted the data, and wrote the manuscript. **PB** assisted in performing the experiments. **DB** helped in FACS analysis and determination of TPO peptides. **PD** and **RW** helped in design of study, recruited and supervised the patient selection and management. **NS** conceptualized and designed the study, supervised the experiments and edited the manuscript.

## Abstract

**Background:** Hashimoto’s thyroiditis (HT) involves the autoimmune destruction of thyrocytes with the presence of anti-thyroid peroxidase (TPO) and/or anti-thyroglobulin (TG) antibodies. In autoimmune diseases, an immunomodulatory role of Bacillus Calmette-Guerin (BCG) vaccination has been reported with decreased autoantibody production and induction of regulatory T cells. We hypothesize that a decline in the efficacy of BCG might be associated with the appearance of HT during adulthood.

**Methods:** Adult subjects with subclinical HT (HT-SCH) (n=39) and non-autoimmune subclinical hypothyroidism (NA-SCH) (n=25) were enrolled along with euthyroid healthy controls (HC) (n=20). The BCG-specific immune responses were determined by the Mantoux test and BCG-induced in-vitro proliferation of peripheral blood mononuclear cells (PBMC). Anti-thyroid cellular immune responses were assessed in subjects with human leukocyte antigen (HLA)-A*02 and HLA-A*24 alleles by determining the frequency of CD8+ T cells recognizing major histocompatibility complex (MHC) dextramers (DMR) carrying TPO-derived peptides, by flow cytometry.

**Results:** The HT-SCH group had a lower rate of Mantoux test reactivity (38.5%) as compared to NA-SCH (64%) and HC subjects (65%) (p=0.047). However, the BCG-induced in-vitro proliferation of PBMC was similar in HT-SCH, NA-SCH, and HC groups. Further, the BCG-vaccinated SCH subjects had lower levels of thyroid stimulating hormone (TSH) (p=0.026). Next, the SCH subjects had higher frequencies of peripheral DMR+ CD8+ T cells as compared to HC subjects (p=0.001). Interestingly, the frequency of peripheral DMR+ CD8+ T cells was significantly higher in HT-SCH than HC subjects (p=0.045). Finally, we observed a positive correlation between the frequency of DMR+CD8+ T cells and TSH levels (r = +0.620, p=0.006).

**Conclusions:** Collectively, our results highlight a complex relationship of neonatal BCG vaccination with the genesis of HT, via modulation of autoimmune responses directed towards thyroid autoantigens.

## INTRODUCTION

Hashimoto’s thyroiditis (HT) is an autoimmune disorder of the thyroid gland characterized by lymphocytic infiltration and destruction of thyrocytes. The prevalence of HT varies from 5-10% and is on the rise, with female preponderance and advancing age [1–4]. The initial pathogenesis of HT involves underlying genetic susceptibility and an environmental trigger like infection, stress, etc., and is seen commonly in iodine-sufficient areas [5]. Various genes implicated in the pathogenesis of HT include human leucocyte antigen (HLA-DR3, HLA-A2, HLA-A24), cytotoxic T lymphocyte-associated protein-4 (CTLA-4), and protein tyrosine phosphatase-22 (PTPN22), etc. [6,7]. Further, there is systemic interaction between T cells, antigen-presenting cells (APCs), and thyroid cells [8,9]. The pathological changes in HT appear with lymphocytic infiltration causing gradual destruction of thyroid follicles, followed by the replacement of thyroid parenchyma with fibrous tissue labeled as atrophic thyroiditis [10].

Bacillus Calmette Guérin (BCG) vaccine is administered during the neonatal stage in countries where tuberculosis (TB) is endemic. BCG has a protective effect against disseminated TB and meningitis in children, however, in adults, the immunoprotection against pulmonary TB declines due to a gradual loss in its efficacy [11,12]. In addition, an immunomodulatory role of BCG associated with a decrease in autoantibodies, and induction of regulatory T cells (Tregs) has been suggested [13]. BCG has also been shown to halt the pathogenesis of other autoimmune diseases, type-1 diabetes (T1D) and multiple sclerosis (MS) by promoting the apoptosis of activated CD4+ T cells [14–17]. Since HT is a manifestation that appears mainly in adulthood, it could be possible that loss of efficacy of BCG might be associated with the appearance of thyroiditis. In a previous report, the BCG vaccine did not prevent the pathogenic initiation of autoimmunity against thyroid-stimulating hormone Receptor (TSHR) but halted the progression to Graves’ disease [18,19]. However, the exact mechanisms behind this immunomodulation are still elusive. To date, there is no definitive evidence in humans on the protective or therapeutic effects of BCG in HT. In this direction, our study is aimed to understand, whether, the onset of HT is associated with a decline in the efficacy of BCG vaccination. Here, we assessed BCG-specific immune responses in the context of anti-thyroid autoimmune responses in recently diagnosed subclinical HT in comparison to non-autoimmune subclinical hypothyroid (NA-SCH) and euthyroid healthy control (HC) subjects.

This article was previously presented as a meeting abstract at the Annual Endocrine Society Conference (Virtual), March 2021 (Session: Thyroid Autoimmunity, COVID-19 & Thyroid Disease. Abstract Number: 7318).

## MATERIALS AND METHODS

### Research design and recruitment of subjects

A total of 84 subjects, aged 18-60 years, presenting with sub-clinical hypothyroidism (SCH) (n=64) and euthyroid normal healthy controls (HC) (n=20) attending departments of Endocrinology at the Post Graduate Institute of Medical Education and Research (PGIMER), Chandigarh, India were enrolled from Oct 2019 to Jan 2021. The SCH subjects were further divided into Hashimoto’s thyroiditis (HT-SCH) (n=39) and non-autoimmune (NA-SCH) (n=25) groups based on the presence of anti-thyroid antibodies. After obtaining informed consent, subjects were screened for TSH, thyroxine (T4), triiodothyronine (T3), free T4, free T3, thyroglobulin (TG), anti-thyroperoxidase (anti-TPO), anti-thyroglobulin (anti-TG) and TSH receptor antibody titers. The criterion for inclusion of SCH subjects was; TSH levels between 4.2-10.0 µIU/ml with normal fT4 levels. Subjects with prior levothyroxine treatment, symptomatic hypothyroidism, infertility, pregnancy or planning for pregnancy, immune deficiency, diabetes mellitus, human immunodeficiency virus (HIV) infection, chronic steroid or immunosuppressant medication, concurrent or past chemotherapy or radiotherapy, active or past tuberculosis, malignancy, and hepatic, renal or cardiac failure were excluded. The study was approved by the institutional ethics committee.

The persistence of the efficacy of BCG vaccination was measured primarily in terms of response to the Mantoux test, and secondly by the presence of a scar formed during primary vaccination (BCG scar). Each subject’s arms and legs were examined for the presence of a BCG scar. For the Mantoux test, 0.1 ml of five tuberculin units (TU) of purified protein derivative (PPD) solution was injected intra-dermally on the volar surface of the upper third region of the forearm by an expert vaccinator in all subjects and examined after 48 hours for induration.

### Peripheral blood mononuclear cell (PBMC) isolation and in-vitro culture with BCG

A total of 10^6^ PBMCs/ml labeled with carboxyfluorescein diacetate succinimidyl ester (CFSE) (5 mM) were plated in triplicates in flat bottom 24-well tissue culture plates (BD-Falcon, USA) in Roswell Park Memorial Institute medium (RPMI)-1640 medium supplemented with 0.1% penicillin, streptomycin, 10% fetal calf serum and stimulated with optimized concentration of 30µl BCG (2-8 x 10^6^ bacilli/ml) (Serum institute, India) or 2µl antiCD3, anti-CD28 DynabeadsTM (Invitrogen, USA) (positive control) or without any stimulant (negative control) for 96 h at 37 °C in 5% CO2 [20,21]. Post culture, the cells were centrifuged at 200g for 10 minutes, acquired on a flow cytometer (FACS Lyric), and analyzed using FACS Suite (version 1.3) software (BD Biosciences, USA). At least 10000 events were acquired and proliferation indices were calculated as described previously [22].

### HLA typing of subjects

DNA was extracted from a 1.0 ml buffy coat of blood samples using QIAamp^TM^ DNA Blood Mini Kit (Qiagen, Hiden, Germany). HLA typing of the recruited subjects was carried out by sequence-specific primers-polymerase chain reaction (PCR-SSP) method (Olerup, Stockholm, Sweden). The PCR products were resolved on a 2% agarose gel. HLA type was interpreted using the SCORE^TM^ software (Olerup, Stockholm, Sweden).

### Dextramer synthesis and validation

HLA class I alleles (A*02, A*24) were selected for the synthesis of peptide dextramers (DMR) as these alleles have a higher affinity for auto-antigenic thyroid peptides and contribute to the development of HT [6]. Two different allophycocyanin (APC) labeled MHC-I DMRs loaded with 8-11mer TPO-derived peptides (A*0201/ALARWLPV, A*2402/VFSTAARF) having the highest cumulative affinity, as determined by MHC-I peptide binding prediction software, Bioinformatics and Molecular Analysis Section (BIMAS), (Centre for Information Technology, NIH, USA), SYFPEITHI (Biomedical Informatics, Heidelberg, Germany) and T cell Epitope Prediction Tool of Immune Epitope Database (IEDB) (La Jolla, CA) were custom synthesized (Immudex, Denmark).

### Immunophenotyping and analysis of TPO-specific CD8+ T cells

The frequencies of TPO-dextramer (DMR) recognizing CD8+ T cells were measured to assess the degree of cellular immune responses against the thyroid gland. After determining HLA-I type, a fresh sample of heparinized peripheral blood was obtained from 14 SCH and 4 HC subjects for immunophenotyping. PBMCs were isolated by density gradient centrifugation with Ficoll (Sigma, USA). After checking cell viability by 7-amino actinomycin D (7-AAD) exclusion, TPO-specific CD8+ T cells were stained by incubating 2 × 10^6^ PBMCs with 10 µl of respective DMRs at room temperature in the dark for 10 min. Anti-CD3 peridinin-chlorophyll (PerCP), anti-CD8 Alexa-fluor 700, and anti-CD4 fluorescein isothiocyanate (FITC) antibodies were then added with 15 min incubation, followed by washing with FACS^TM^ buffer (BD Biosciences, USA). The stained cells were acquired on a flow cytometer (FACS Lyric) and analyzed using FACS Suite (version 1.3) software (BD Biosciences, USA). At least one million total events were acquired and a minimum of 500,000 T cells were analyzed. Gating was first performed on lymphocytes followed by gating of CD3+ CD4+ CD8+ cells to exclude CD3- T cells, monocytes, NK cells, and B cells. CD8+ T cells recognizing the DMRs were then gated and analyzed. Relevant fluorescence minus one (FMO) tubes were used to set gates.

### Statistical Analysis

Statistical analyses were performed using Statistical Package for The Social Sciences (SPSS v22.0) (SPSS Inc., Chicago, IL). Continuous variable data was expressed as mean± Standard Deviation (SD) or median with inter-quartile range (1st – 3rd) and discrete variables as proportions. The student’s t-test or one-way analysis of variance (ANOVA) was used to compare the difference between means of normally distributed variables, while Mann Whitney or Kruskal Wallis was used to compare the difference between means of variable with inequality of variance. The chi-square test or Fischer exact was used to examine the difference between proportions. Comparison between the three groups was done using Tukey post hoc analysis. The Pearson correlation coefficient was used for the correlation of parametric data, whereas Spearman correlation analysis was performed for non-parametric data. All comparisons were done at a level of significance of 0.05.

## RESULTS

### Clinical characteristics of the study subjects

Of the total 64 subclinical hypothyroid subjects, 39 subjects had subclinical Hashimoto’s thyroiditis (HT-SCH), 25 subjects had non-autoimmune SCH (NA-SCH) and 20 subjects were non-autoimmune euthyroid healthy controls (HC). The clinical and biochemical characteristics including TSH, T4, T3, fT4, and fT3 are documented in Table 1. The thyroid profile was repeated for all subjects at 6 months as a part of regular work-up and the intra-subject thyroid profile and anti-thyroid antibody status was similar at both time points. Other biochemical characteristics of the subjects are shown in Table 2.

**Table 1:** Clinical and biochemical characteristics of HT-SCH, NA-SCH and HC subjects.

| Variables | HT-SCH (n=39) | NA-SCH (n=25) | HC (n=20) | P-value |
| --- | --- | --- | --- | --- |
| Age (years)* | 37.64±11.5 | 34.92 ±10.3 | 33.5±8.02 | 0.313 |
| Sex (Female), n (%) | 33 (84.6) | 16 (64) | 16 (80) | 0.149 |
| BMI (kg/m<sup>2</sup>) | 25.8±4.65 | 25.4±7.09 | 25.66±3.20 | 0.944 |
| BCG scar, Present, n (%) | 34 (87.18) | 19 (76) | 19 (95) | 0.182 |
| TSH (μIU/ml)* | 7.0±1.7 | 6.8±1.51 | 2.3±0.66 | 0.001<sup>+</sup> |
| fT4 (ng/dl)* | 1.34±0.35 | 1.43±0.41 | 1.22±0.33 | 0.124 |
| fT3 (pmol/L)* | 5.39±0.94 | 5.87±1.15 | 5.45±0.42 | 0.159 |
| Anti TG Ab (IU/ml)<sup>#</sup> | 72.5 (11.7-59.6) | 5 (5-9.8) | 5 (5-9.0) | 0.001<sup>++</sup> |
| Anti-TPO Ab (IU/ml)<sup>#</sup> | 294.6 (120.2-764.2) | 26.4 (14.1-47.44) | 10 (10-14.3) | 0.001<sup>++</sup> |

\* Mean (±SD), P-value was calculated by one-way ANOVA test for parametric data, # Median (1<sup>st</sup> - 3<sup>rd</sup> quartile), P-value was calculated by Kruskal Wallis test for continuous non-parametric data and Chi-square test for categorical data. Abbreviations: HT-SCH Subclinical Hashimoto's thyroiditis, NA-SCH: Non-autoimmune subclinical hypothyroidism, HC: Healthy control, BMI: Body mass index, BCG: Bacillus Calmette-Guerin, TSH: Thyroid stimulating hormone, fT4: Free thyroxine, fT3: Free triiodothyronine, TG: Thyroglobulin, TPO: Thyroid peroxidase, Ab: Antibody; (+) p-value between HT-SCH and NA-SCH vs. HC, (++) p-value between HT-SCH vs. HC and NA-SCH subjects

**Table 2:** Other biochemical characteristics of HT-SCH, NA-SCH and HC subjects.

| Variables | HT-SCH<br>(n=39) | NA-SCH (n =<br>25) | HC (n = 20) | P-value |
| --- | --- | --- | --- | --- |
| Family History of Hypothyroidism, Present, n (%) | 12 (31%) | 4 (16%) | 5 (25%) | 0.390 |
| Thyroglobulin (ng/ml) | 8.95 (1.7-43.22) | 16.65 (7.63-25.0) | 16.7 (11.5-22.1) | 0.413 |
| CRP (mg/L) <sup>#</sup> (Normal= 0-6) | 2.44 (0.87-5.2) | 3.12 (1.9-5.6) | 3.1 (1-5.5) | 0.810 |
| Vitamin B12 (pg./ml) <sup>#</sup> | 219.7 (144-360) | 215 (125.8-391) | 256 (155-449) | 0.638 |
| Iron deficiency, n (%) | 16 (41%) | 10 (40%) | 6 (28.6%) | 0.653 |
| Vitamin D (ng/ml) <sup>#</sup> | 15.7 (9.9-22.9) | 16.23 (10-24.7) | 12 (7-20.1) | 0.328 |
| Total cholesterol (mg/dl)* | 169.13±27.45 | 174.97±41.36 | 150.46±27.13 | 0.043<sup>+</sup> |
| LDL (mg/dl)* | 106.61±24.67 | 109.38±36.40 | 88.94±25.56 | 0.056 |
| HDL (mg/dl)* | 45.43±11.91 | 51.07±9.26 | 45.03±8.84 | 0.234 |
| Triglyceride (mg/dl)* | 129.13±63.93 | 116.82±56.53 | 124.21±51.22 | 0.749 |
| Hemoglobin (g/dl)* | 12.24±1.30 | 12.35±1.9 | 12.49±1.37 | 0.819 |
| Urea (mg/dl)* | 20.64±6.36 | 21.86±5.46 | 18.64±6.8 | 0.157 |
| Uric acid (mg/dl)* | 4.44±1.09 | 4.69±1.07 | 3.69±1.3 | 0.705 |
| Creatinine (mg/dl)* | 0.74±0.24 | 0.74±0.15 | 0.68±0.19 | 0.467 |
| Total protein (g/dl)* | 7.52±0.44 | 7.55±0.57 | 7.53±0.52 | 0.979 |
| Albumin (g/dl)* | 4.46±0.33 | 4.51±0.41 | 4.33±0.37 | 0.174 |
| Bilirubin (mg/dl)* | 0.57±0.25 | 0.60±0.26 | 0.73±0.27 | 0.066 |
| ALP (U/L) * | 94.47±40.5 | 89.48±21.19 | 81.45±22.93 | 0.313 |
| AST (U/L)* | 27.35±7.48 | 25.97±14.69 | 27.6±8.8 | 0.852 |
| ALT (U/L)* | 28.53±14.63 | 31.90±27.59 | 27.65±17.51 | 0.723 |

\* Mean(±SD), P-value was calculated by one-way ANOVA test for parametric data, # Median (1st - 3rd quartile), P-value was calculated by Kruskal Wallis test for continuous non- parametric data, and Chi-square test for categorical data. (+) p-value between NA-SCH and HC subjects. Abbreviations: HT-SCH Subclinical Hashimoto's thyroiditis, NA-SCH: Non- autoimmune subclinical hypothyroidism, HC: Healthy control, CRP: C-reactive protein, LDL: Low-density lipoprotein, HDL: High-density lipoprotein, ALP: Alkaline phosphatase, AST: aspartate aminotransferase, ALT: Alanine transaminase.

### Assessment of efficacy of primary BCG vaccination

The efficacy of primary (neonatal) BCG vaccination was determined by Mantoux test reactivity and by in-vitro proliferative responses of peripheral blood lymphocytes to BCG. The Mantoux test-associated induration was observed in fewer HT-SCH subjects (induration diameter ≥ 5mm) (38.5%) as compared to NA-SCH (64%) and HC (65%) subjects (p=0.047) (Figure 1) (Table 3), with odds ratio (OR) between HT-SCH versus HC and NA-SCH subjects being, 0.34 (0.11-1.0, p=0.05) and 0.35 (0.12-0.99, p=0.046), respectively. Secondly, the peripheral blood mononuclear cells were isolated from the subjects and cultured in the presence of BCG vaccine (Figure 2). In contrast to the Mantoux test, the in-vitro lymphocyte proliferation in the presence of BCG was similar in HT-SCH and NA-SCH subjects (proliferation index, 2.55±0.31 vs. 2.51±0.41, p=0.889).

**Figure 1:**
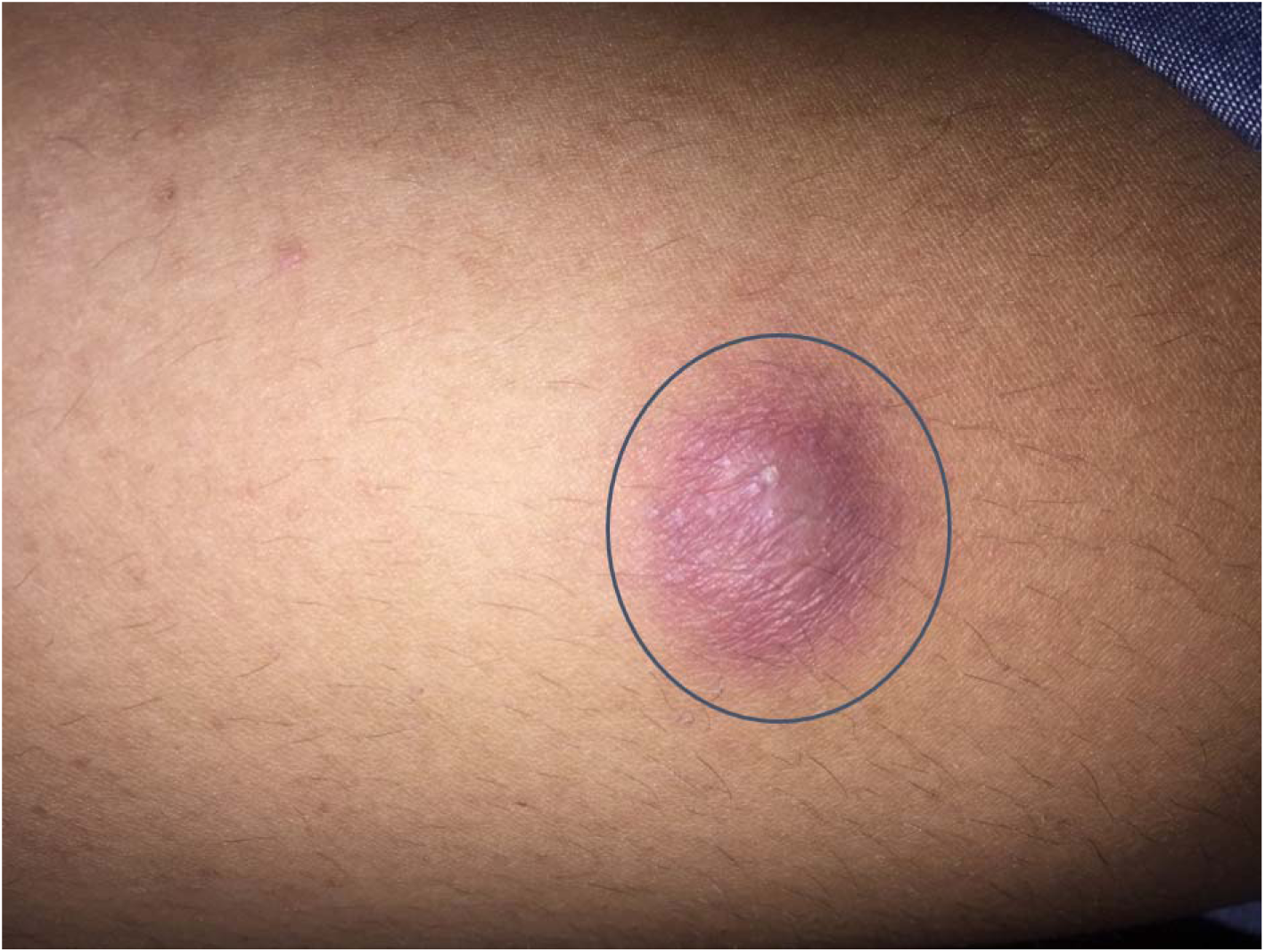
Representative image of induration at forearm, after 48 hours of Mantoux test. **Legend:** An induration diameter < 5mm was considered as non-reactive.

**Figure 2:**
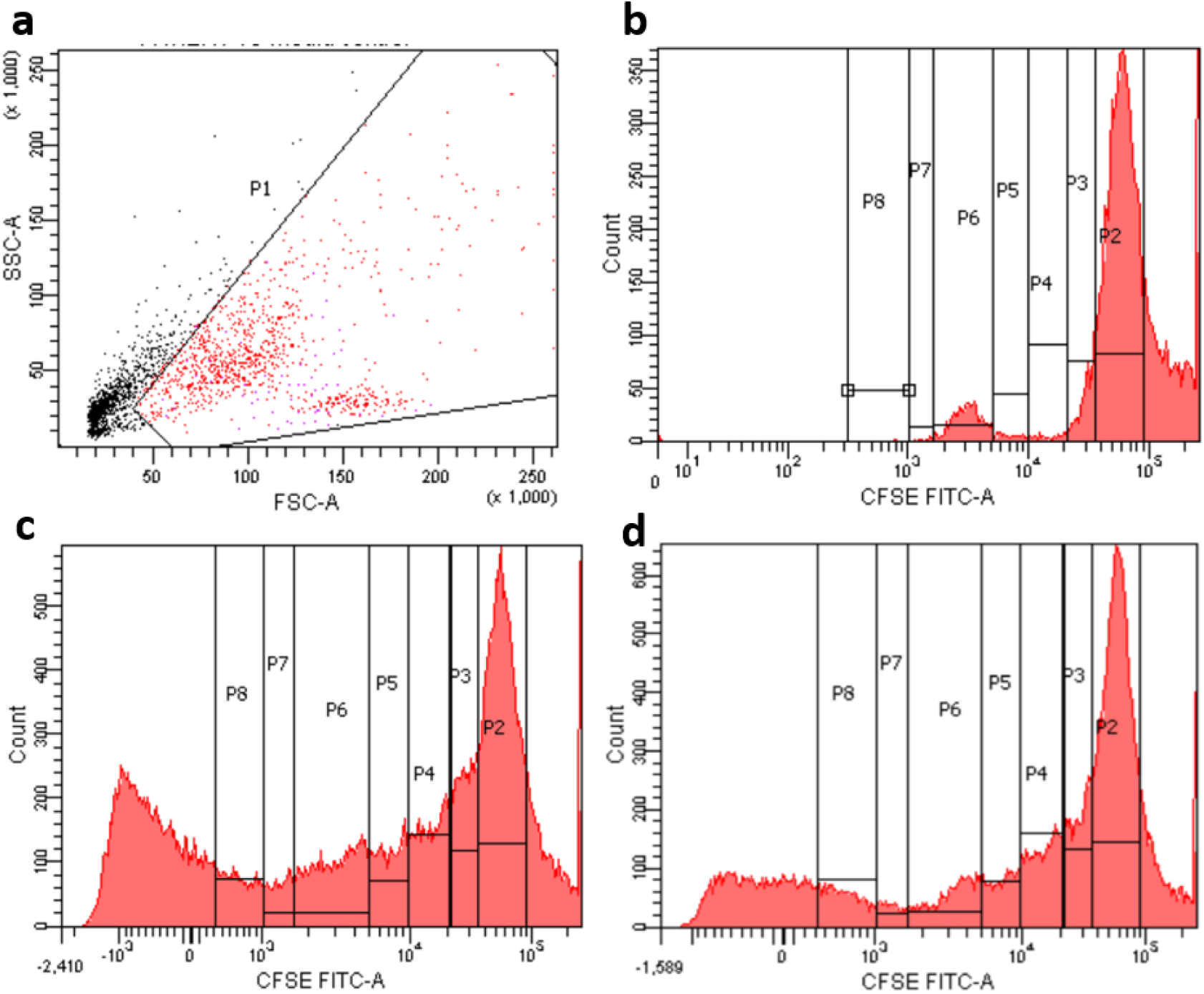
Representative flow-cytograms showing proliferation of CFSE labelled peripheral blood mononuclear cells (PBMC) isolated from the peripheral blood of a subject with subclinical Hashimoto’s thyroiditis (HT-SCH). **Legend:** The PBMCs were labeled with CFSE (5mM) at RT for 20 min then stimulated in-vitro with BCG or anti-CD3, anti-CD28 Dynabeads for 96 hours and analyzed on a flow cytometer. P2 represents the parent population, whereas P3-P8 represent the daughter population after consecutive divisions. a) Lymphocytes were gated as per forward and side scatter profiles. The proliferation of CFSE labeled lymphocytes in, b) unstimulated cultures (negative control, MC), (c), in the presence of anti-CD3, anti-CD28 Dynabeads^TM^ (positive control), and (d), in the presence of BCG (30 µl/well).

**Table 3:** Mantoux test response and BCG-induced *in vitro* lymphocyte proliferation in HT-SCH, NA-SCH and HC subjects.

| Outcome variables |  | HT-SCH<br>(n=39) | NA-SCH<br>(n=25) | HC (n=20) | P-value |
| --- | --- | --- | --- | --- | --- |
| Mantoux | Positive, n (%) | 15 (38.5) | 16 (64) | 13 (65) | 0.047<sup>+</sup> |

| Test | Negative, n (%) | 24 (61.5) | 09 (36) | 07 (35) |  |
| --- | --- | --- | --- | --- | --- |
| Proliferation index (BCG)* |  | 2.55±0.31 | 2.51±0.41 | 2.46±0.29 | 0.667 |
| Proliferation index (anti-CD3/CD28) * |  | 2.59±0.26 | 2.53±0.39 | 2.44±0.26 | 0.285 |
| Proliferation index (MC)* |  | 2.51±0.28 | 2.44±0.38 | 2.40±0.27 | 0.677 |

\* Mean(±SD) p-value was calculated by independent-samples t-test for parametric data and chi-square test for categorical variables. (+) p-value between HT-SCH vs. NA-SCH and HC subjects. Abbreviations: HT-SCH: subclinical Hashimoto's thyroiditis, NA-SCH: non-autoimmune subclinical hypothyroidism, HC: healthy control, BCG: Bacillus Calmette-Guerin, CD: cluster of differentiation, MC: Media control.

### Assessment of autoimmune responses to thyroid antigens

As a measure of humoral autoimmunity towards the thyroid gland, we assessed the titers of anti-TPO, anti-TG, and anti-TSHR antibodies in all subjects. Amongst the SCH group, anti-TPO, anti-TG, and both (anti-TPO and anti-TG) antibodies were positive in 64%, 77%, and 41% of subjects respectively. The levels of anti-TSHR antibodies were low and well below the cut-off limit (<0.8 IU/L) in all three groups, indicating the absence of humoral immune responses toward TSHR.

To determine the HLA-specific cellular immune responses toward the thyroid gland, we performed HLA-typing of the subjects and chose TPO as the target antigen. At first, we observed heterogeneity of HLA-I alleles in our study subjects. HLA typing was performed in 52/84 subjects, including 41 SCH and 11 healthy controls. In the case of MHC-I, HLA-A*24 (41%) was the most frequent allele in SCH subjects followed by HLA-B*07 (37%) and HLA-A*01 (31%). HLA-A*02 was seen in 27% of subjects of SCH. In the case of MHC class II, the most frequent allele was HLA-DQB1*02 (78%) followed by HLA-DRB1*01 (65%) (Figure 3). Based on the frequency of HLA-I alleles, we assessed the frequency of peripheral CD8+ T cells that bind to TPO-derived epitopes using MHC-I dextramers. These autoreactive cells were termed DMR+ CD8+ T cells and analyzed by flow cytometry (Figure 4).

**Figure 3:**
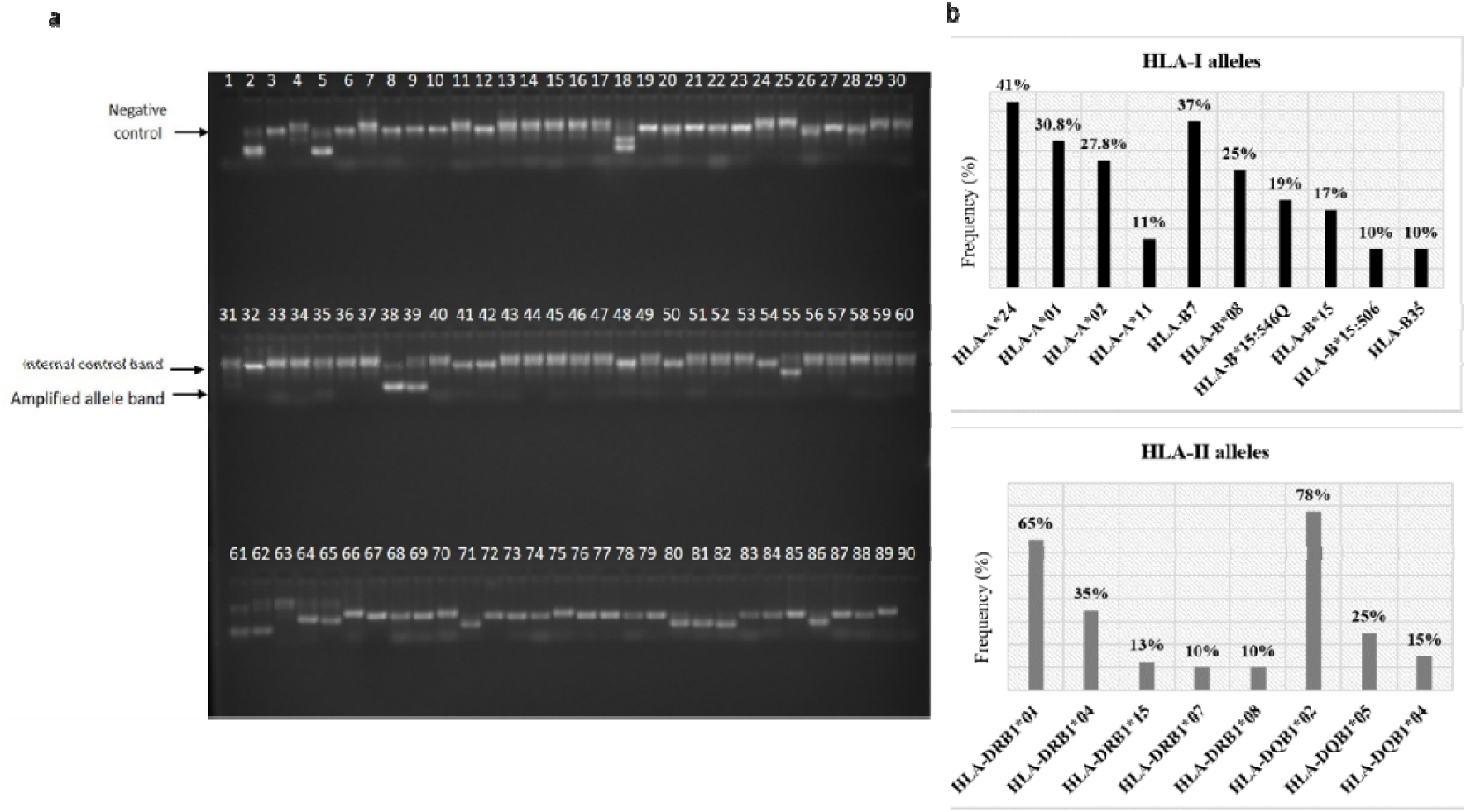
HLA alleles of the study subjects. **Legend: a:** A representative agarose gel image of PCR amplified products during molecular HLA typing (PCR-SSP), showing the internal control band, amplified allele band and negative control. Lane 1 represents negative control. The amplified internal control is present in all the lanes except negative control. Amplified allele bands are observed in lanes 2, 5, 18, 38, 39, 55, 1, 62, 64, 65, 71, 80, 81, 82 and 86. HLA type of the subjects was determined using SCORE^TM^ software provided by the manufacturer (Olerup, Stockholm, Sweden). **b:** Frequency of various HLA I and HLA II alleles determined by molecular HLA typing in SCH subjects.

**Figure 4:**
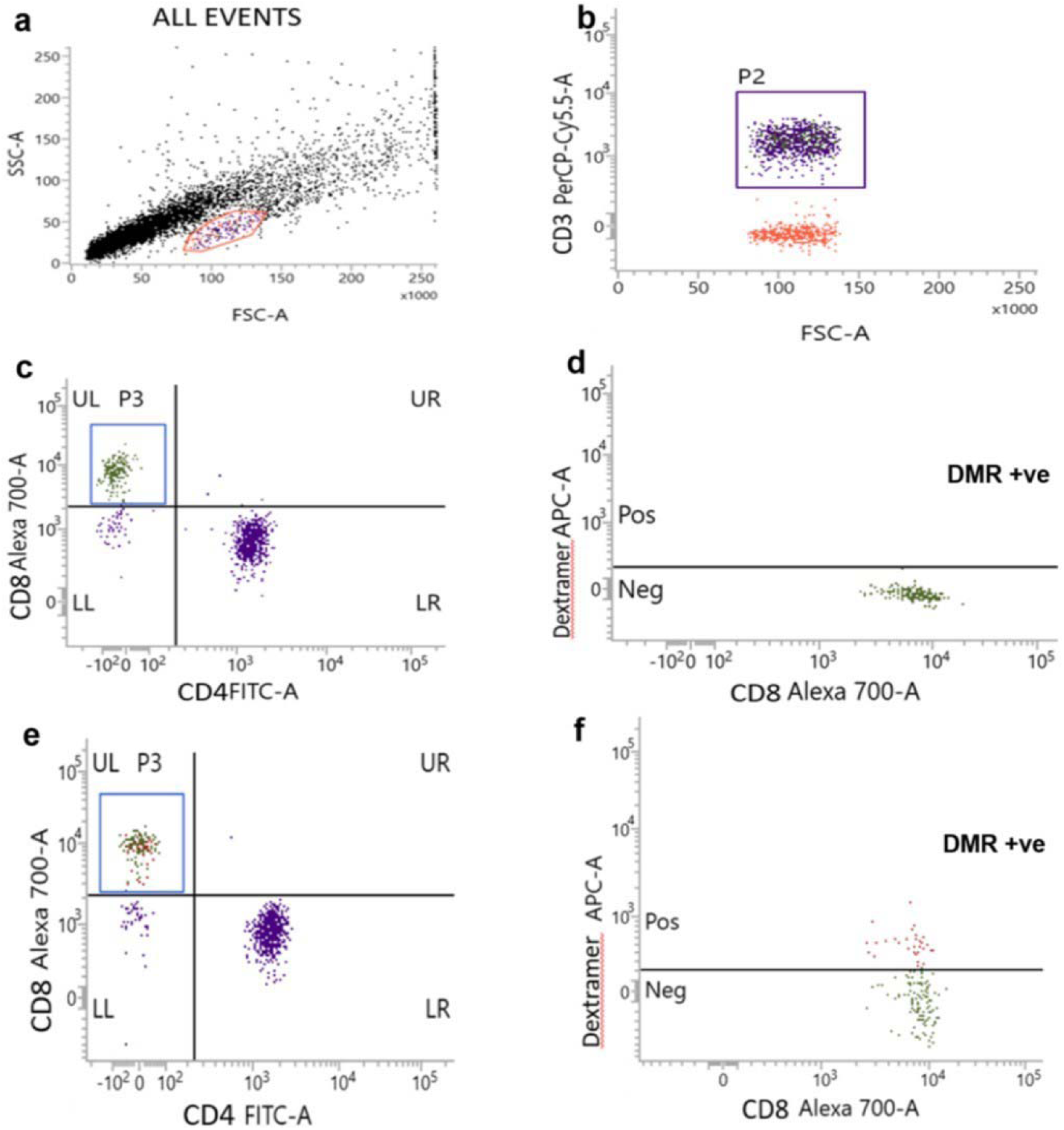
Representative images showing gating strategy for the immunophenotyping of TPO-specific autoreactive CD8+ T (DMR+ CD8+ T cells) cells in the peripheral blood from a HT-SCH subject. **Legend:** a) Lymphocytes were gated as per forward and side scatter profiles, b) gating of CD3+ T cells, c) gating of CD8+CD4- T cells in FMO tube, d) DMR+CD8+ T cells set in FMO tube for negative control, e) gating of CD8+CD4- T cells and f) CD8+ T cells recognizing DMRs carrying TPO derived peptide selected as DMR+ve cells.

We took HLA-A*02 and HLA-A*24 negative HT-SCH subjects as negative controls to determine the frequency of DMR+CD8+ T cells in SCH subjects in comparison to HC subjects as described previously [20]. The average frequency (%) of DMR+CD8+ T cells was deducted from the frequency (%) of DMR+CD8+ T cells in HLA-A*02 and HLA-A*24 positive subjects to calculate the actual frequency (%) of autoreactive CD8+ T cells recognizing TPO epitopes. At first, to rule out whether the SCH subjects had higher frequency of peripheral CD8+ T cells, we calculated the CD4:CD8 T cell ratios and observed that HC and SCH subjects had similar CD4: CD8 T cell ratios (2.57±1.1 vs. 2.06±1.09, p=0.438), Interestingly, we observed that the SCH subjects had higher frequencies of DMR+ CD8+ T cells (7.6±4.46% vs. 1.61±0.80%, p=0.001) as compared to healthy controls.

The calculated frequency (%) of DMR+ CD8+ T cells was found to be similar in NA-SCH and HT-SCH subjects [mean (±SD) 6.6±4.33% vs. 9.02±4.17%, p=0.737]. As expected, the DMR+ CD8+ T cells were significantly higher in HT-SCH than in HC subjects (9.02±4.17 vs. 1.61±0.80 %, p=0.045) (Table 4) (Figure 5).

**Figure 5:**
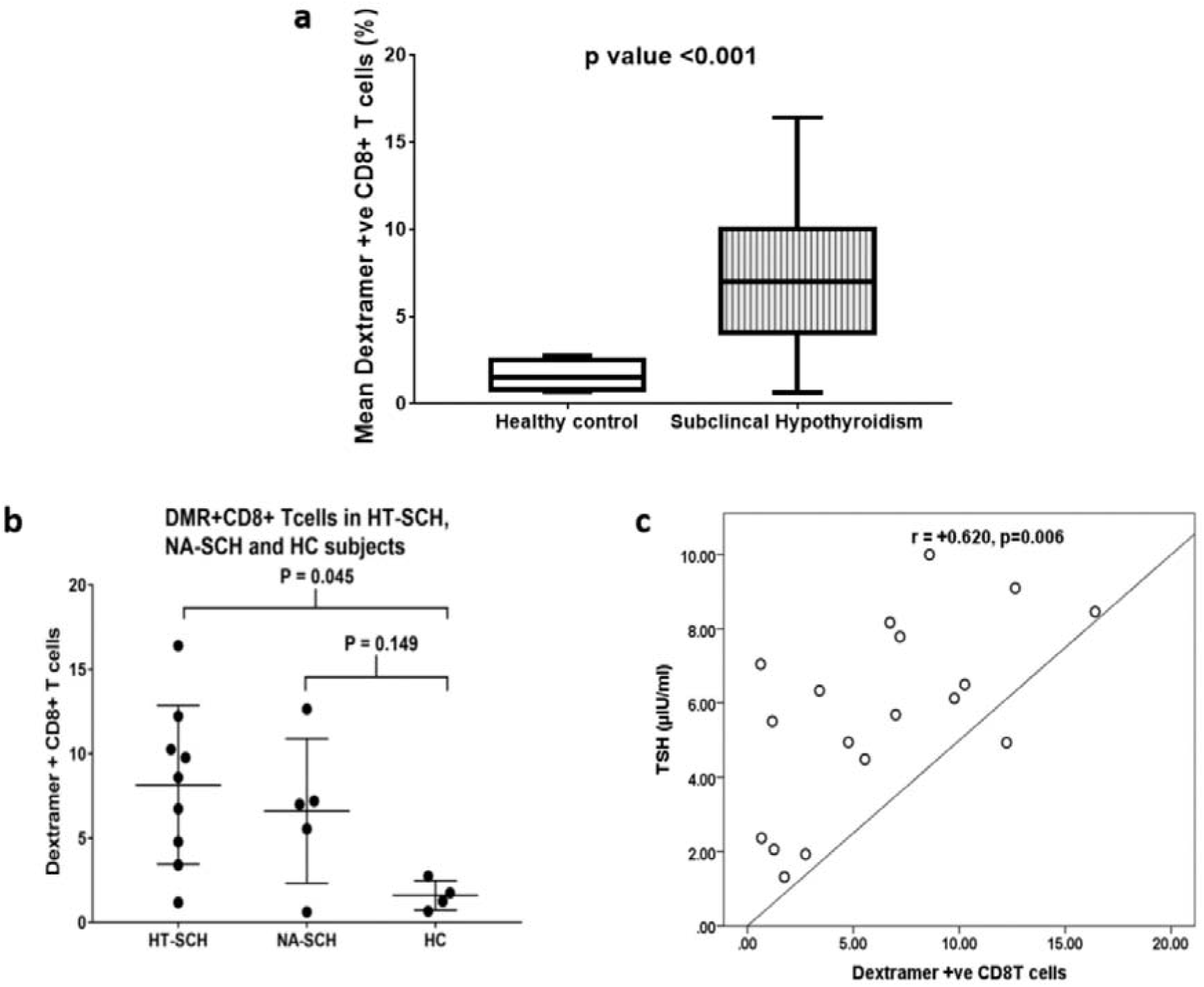
Assessment of anti-thyroid autoreactive CD8+ T cell responses. **Legend:** a) Comparison of the mean relative frequency of DMR+ CD8+ T cells in the peripheral blood of subclinical hypothyroid (SCH) and healthy control (HC) subjects. b) Comparison of the mean relative frequency of DMR+CD8+ T cells in subclinical Hashimoto’s thyroiditis (HT-SCH), non-autoimmune SCH (NA-SCH) subjects, and healthy controls (HC). c) Correlation between TSH levels and DMR+CD8+ T cells (amongst CD8+ T cells). Data is presented as mean±SD and Student’s t-test was used to compare the means in figure (a) whereas, one-way ANOVA followed by Tukey’s multiple comparison test was used to compare the means in figure (b), and correlation is presented as Pearson coefficient in figure (c). P < 0.05 was considered as statistically significant.

**Table 4:** Cellular autoimmune responses to thyroid antigens in SCH and HC subjects.

| Outcome variables | HT-SCH<br>(n=09) | NA-SCH<br>(n=05) | HC (n= 04) | P-value |
| --- | --- | --- | --- | --- |
| CD3+ Lymphocytes (%)* | $60.28 \pm 8.53$ | $68.12 \pm 3.55$ | $65.0 \pm 15.97$ | 0.336 |
| CD3+CD8+ T cells (%)* | $18.47 \pm 7.32$ | $21.67 \pm 6.43$ | $21.17 \pm 14.90$ | 0.723 |
| CD3+CD4+ T cells (%)* | $35.71 \pm 10.01$ | $36.56 \pm 7.64$ | $42.68 \pm 16.65$ | 0.559 |
| CD4: CD8 T cell ratio* | $2.18 \pm 1.01$ | $1.83 \pm 0.74$ | $2.57 \pm 1.1$ | 0.555 |
| DMR +ve (CD8+ T cells) (%)* | $9.02 \pm 4.17$ | $6.6 \pm 4.33$ | $1.61 \pm 0.80$ | $0.045^+$ |

\* Mean ( ± SD), P value was calculated by one-way ANOVA test. (+), p-value between HT-SCH vs. HC subjects. Abbreviations: HT-SCH: subclinical Hashimoto's thyroiditis, NA-SCH: non-autoimmune subclinical hypothyroidism, HC: healthy control, DMR: APC labelled MHC-I Dextramers loaded with thyroid peroxidase (TPO) derived peptide, BCG: Bacillus Calmette-Guerin, CD: Cluster of differentiation.

Next, we observed a significant positive correlation between the frequency of DMR+CD8+ T cells and TSH levels (r=+0.620, p=0.006) in the complete cohort, indicating that the appearance of TPO-specific CD8+ T cells is associated with the onset of hypothyroid disease. However, there was no significant correlation between the frequency of DMR+CD8+ T cells and anti-TPO antibody titers (r=+0.207, p=0.409) or anti-TG antibody titers (r=+0.108, p=0.669).

### Comparison of SCH subjects based on BCG vaccination

The presence or absence of a scar at the site of BCG administration may suggest, whether the subject has been vaccinated (Table 5). Overall, the TSH levels were significantly lower in BCG-vaccinated subjects (presence of a BCG scar) (6.75±1.56 vs. 7.94±1.67 µIU/ml, p=0.026) as compared to non-BCG vaccinated subjects (absence of a BCG scar). The non-BCG-vaccinated SCH subjects had a lower BMI (22.15±2.81 vs. 26.34±5.82, p=0.030) as compared to the BCG-vaccinated SCH subjects. Overall, the anti-mycobacterial response i.e., Mantoux test (induration in mm) was comparable in both the groups (p=0.663). Also, the non-BCG vaccinated SCH subjects had similar in-vitro BCG-induced lymphocyte proliferation response (2.38±0.29 vs. 2.56±0.36, p=0.127) as compared to the BCG vaccinated SCH subjects.

**Table 5:** Thyroid profile, anti-mycobacterial responses and BCG-induced *in-vitro* T cell proliferation in SCH subjects with or without BCG vaccination.

| Variables |  | BCG scar absent<br>(n = 11) | BCG scar present<br>(n = 53) | P-value |
| --- | --- | --- | --- | --- |
| Age (years)* | | 31.92±7.75 | 36.20±11.14 | 0.118 |
| BMI (kg/m<sup>2</sup>)* | | 22.15±2.81 | 26.34±5.82 | 0.030 |
| TSH (μIU/ml)* | | 7.94±1.67 | 6.75±1.56 | 0.026 |
| fT4(ng/dl)* | | 1.52±0.55 | 1.34±0.33 | 0.376 |
| fT3(pmol/L)* | | 5.74±0.92 | 5.53±1.07 | 0.468 |
| Anti –TPO Ab (IU/ml)<sup>#</sup> |  | 12.0 (5-150) | 16.4 (5-75) | 0.913 |
| Anti - TG Ab (IU/ml)<sup>#</sup> |  | 56.0 (11-454) | 106.9 (37.3-345.9) | 0.516 |
| Mantoux<br>Test | Negative n (%) | 5 (45.45) | 28 (52.83) | 0.663 |
|  | Positive n (%) | 6 (54.55) | 25 (47.17) |  |
| Proliferation index (BCG)* | | 2.38±0.29 | 2.56±0.36 | 0.127 |

\* Mean( ± SD), the p-value was calculated by independent-samples t-test for parametric variables. # Median (1st – 3rd quartile), the p-value was calculated by Mann Whitney U test for continuous non-parametric data and Chi-square test for categorical data. Abbreviations: BMI: Body mass index, BCG: Bacillus Calmette-Guerin, TSH: Thyroid stimulating hormone, fT4: Free thyroxine, fT3: Free triiodothyronine, TG: Thyroglobulin, TPO: Thyroid peroxidase, Ab: Antibody. The subjects with the presence of a BCG scar were considered as BCG vaccinated.

## DISCUSSION

Hashimoto’s thyroiditis is primarily mediated by autoreactive T cells that induce the destruction of thyrocytes. BCG vaccine has been shown to have an immunomodulatory role in autoimmune diseases like T1DM, and MS [15,16,23], however, to the best of our knowledge, there is no human study on the role of BCG vaccination in HT or related autoimmune thyroid diseases. We included only SCH subjects and attempted to assess the effects of prior BCG vaccination on anti-thyroid immune responses and its impact during the early pathogenesis of the disease

Overall, most of the HT-SCH subjects were non-reactive to the Mantoux test as compared to NA-SCH and HC subjects. Amongst the SCH subjects, the BCG-vaccinated group showed lower levels of TSH in comparison to the non-vaccinated group. Additionally, those SCH subjects retaining the efficacy of BCG had almost significantly lower titers of anti-TG antibodies. Together, these observations indicate that the persistence of BCG efficacy has some role in regulating autoimmune responses towards the thyroid gland. It is also pertinent to mention here that the BCG scar may be absent in 1-20 % of subjects even after vaccination [24,25]. There are a few reports where BCG vaccination has been shown to be associated with a lower frequency of autoreactive T cells resulting in a decrease in the pathogenesis of autoimmune diseases [14]. In the case of thyroid, in a mouse model of Graves’ disease, Nagayama et al. (2004), demonstrated that BCG vaccination slows disease progression by diverting the anti-TSHR directed Th1 cells and reducing the production of anti-TSHR antibodies [19]. In another study, GAD65 and IA-2 autoantibody titers were observed to be lower in BCG-vaccinated (ascertained by the presence of a BCG scar) T1D subjects [26]. Likewise, in our study, prior BCG vaccination was associated with lower levels of TSH in SCH subjects.

In contrast to Mantoux reactivity, we did not find any difference between in-vitro BCG-induced PBMC proliferation in subjects in the context of their BCG vaccination status. We think that there could be inherent potential differences between in-vivo and in-vitro responses of T cells to BCG and this assay might not ascertain BCG vaccination efficacy, efficiently, particularly in the Indian subcontinent due to the likelihood of prior exposure to subclinical/environmental tuberculosis, non-tubercular infections. The absence of correlation between in-vivo and in-vitro BCG-specific immune responses has been seen in similar studies as well. Biswas et al. 1997 observed no correlation between Mantoux response and PPD-induced lymphocyte proliferation in Eales’ disease [27]. In another study, Arya et al. (2018) observed that both healthcare workers and active tuberculosis patients had similar lymphocyte proliferation in response to Mycobacterium tuberculosis (MTB) Memory antigen irrespective of their Mantoux test reactivity [28]. These findings suggest that subclinical/environmental exposure to MTB in areas with a prevalence of tuberculosis influences BCG-associated immune responses.

To study autoreactive T cells, various TPO peptides have been used previously [20]. We could demonstrate peripheral autoreactive CD8+ T cells recognizing, TPO-derived peptides in both HT-SCH and NA-SCH subjects. The presence of autoreactive CD8+ T cells in NA-SCH could be attributed to higher levels of TSH, and without studying their phenotype it would be too early to label these DMR+CD8+ T cells as pathogenic. A similar study reported the presence of autoreactive CD8+ T cells recognizing TG and TPO epitopes in anti-TG and anti-TPO antibody-negative subjects [21]. Therefore, these observations support the occurrence of positive cellular immune responses even in the absence of humoral immune responses toward thyroid antigens [20,21]. However, we cannot comment on whether this could be attributed to the ‘trained immunity’ imparted by BCG, since we did not assess BCG reactive T cells in these subjects.

Another relevant finding from our study was a positive correlation between TSH levels and autoreactive CD8+T cells suggesting initiation of cellular autoimmunity leading to thyroid parenchymal destruction before the onset of humoral responses. Similarly, Ehlers et al. (2012) showed a lack of correlation between anti-TPO Ab and autoreactive CD8+ T cells [20]. This varying correlation of anti-thyroid antibodies with autoreactive CD8+ T cells probably highlights the fact that autoreactive CD8+ T cells emerge earlier than the autoantibody-producing B cells, which likely have a bystander or a surrogate role rather than being pathogenic [6,10,29]. Since cytotoxic CD8+ T cells primarily mediate damage to the thyroid gland, their appearance in both HT-SCH and NA-SCH subjects is an indication that the onset of hypothyroidism perhaps also initiates a phase of autoimmunity. However, a smaller number of subjects available in the non-BCG vaccinated group, lack of objective evidence of iodine sufficiency, and fine needle aspiration cytology (FNAC) of the thyroid gland to confirm the diagnosis of autoimmunity are the main limiting factors in our study. Nevertheless, this is the first human study ever done in HT subjects highlighting the association between appearances of anti-thyroid autoimmunity in the context of BCG vaccination.

## Conclusions

In conclusion, our study highlighted a complex relationship between neonatal BCG vaccination and the appearance of autoimmunity in the thyroid gland in SCH subjects. Our results suggest that BCG vaccination has some influence on the genesis of HT by altering the frequencies of autoreactive T cells and the appearance of antibodies directed toward thyroid autoantigens. BCG is a time-tested safe vaccine that has already shown its non-specific effects in other infections and autoimmune diseases. We have now partially demonstrated that neonatal BCG vaccination exerts an immunomodulatory role on thyroid autoimmunity. Our data warrants further long-term human studies in adult subjects with family history or early onset of HT to assess the utility of BCG in therapeutic vaccination.

## Data Availability

All data produced in the present study are available upon reasonable request to the authors

## Abbreviations

APC: Allophycocyanin
BCG: *Bacillus Calmette–Guérin*
CFSE: Carboxyfluorescein diacetate succinimidyl ester
DMR: Dextramer
7-AAD: 7-aminoactinomycin D
FITC: Fluorescein isothiocyanate
FMO: fluorescence minus one
FNAC: Fine needle aspiration cytology
HC: Healthy control
HIV: Human immunodeficiency virus
HLA: Human leukocyte antigen
HT: Hashimoto’s thyroiditis
IEDB: Immune epitope database
MHC: Major histocompatibility complex
MS: Multiple sclerosis
MTB: *Mycobacterium tuberculosis*
NA-SCH: Non-autoimmune subclinical hypothyroid
PBMC: Peripheral blood mononuclear cells
PCR: Polymerase chain reaction
PerCP: Peridinin-chlorophyll
PPD: Purified protein derivative
RPMI: Roswell Park Memorial Institute
SCH: Sub-clinical hypothyroidism
SD: Standard deviation
T1D: Type-1 diabetes
T3: Triiodothyronine
T4: Thyroxine
TG: Anti-thyroglobulin
TPO: Anti-thyroid peroxidase
Tregs: Regulatory T cells
TSH: Thyroid stimulating hormone
TU: Tuberculin units

## Acknowledgements

We acknowledge the technical staff of the Department of Endocrinology, PGIMER, Chandigarh, including Mr Vivek Sharma, Mudasir Hassen, Pintoo Kumar, Raj Davinder Kaur for their support and assistance in performing all the laboratory investigations related to this study.

